# THE INFLUENCE OF HLA GENOTYPE ON SUSCEPTIBILITY TO, AND SEVERITY OF, COVID-19 INFECTION

**DOI:** 10.1101/2020.12.31.20249081

**Authors:** David J Langton, Stephen C Bourke, Benedicte A Lie, Gabrielle Reiff, Shonali Natu, Rebecca Darlay, John Burn, Carlos Echevarria

**Affiliations:** ExplantLab, The Biosphere, Newcastle Helix, Newcastle-upon-Tyne, NE4 5BX; North Tyneside General Hospital, North Shields, Tyne and Wear, NE29 8NH; Department of Medical Genetics, University of Oslo and Oslo University Hospital; University Hospital of North Tees, Stockton, TS19 8PE; Institute of Human Genetics, Newcastle University, International Centre for Life, NE1 3BZ; Royal Victoria Infirmary, Newcastle upon Tyne, NE1 4LP

## Abstract

**Background:** The impact of COVID-19 varies markedly, not only between individual patients but also between different populations. We hypothesised that differences in human leukocyte antigen (HLA) genes might influence this variation.

**Methods:** Using next generation sequencing, we analysed the class I and class II classical HLA genes of 147 white British patients with variable clinical outcomes. 49 of these patients were admitted to hospital with severe COVID infection. They had no significant pre-existing comorbidities. We compared the results to those obtained from a group of 69 asymptomatic hospital workers who evidence of COVID exposure based on blood antibody testing. Allelic frequencies in both the severe and asymptomatic groups were compared to local and national healthy controls with adjustments made for age and sex. With the inclusion of hospital staff who had reported localised symptoms only (limited to loss of smell/taste, n=13) or systemic symptoms not requiring hospital treatment (n=16), we carried out ordinal logistic regression modelling to determine the relative influence of age, BMI, sex and the presence of specific HLA genes on symptomatology.

**Findings:** We found a significant difference in the allelic frequency of HLA-DRB1*04:01 in the severe patient compared to the asymptomatic staff group (5.1% versus 16.7%, p=0.003 after adjustment for age and sex). There was a significantly lower frequency of the haplotype DQA1*01:01/DQB1*05:01/DRB1*01:01 in the asymptomatic group compared to the background population (p=0.007). Ordinal logistic regression modelling confirmed the significant influence of DRB1*04:01 on the clinical severity of COVID-19 observed in the cohorts.

**Interpretation:** This study provides evidence that patient age, sex, BMI and HLA genotype interact to determine the clinical outcome of COVID-19 infection.

**Research in context:** *Evidence before this study:* HLA genes are implicated in host resistance or susceptibility to a range of pathogens. No studies thus far have compared HLA allele frequencies in patients requiring hospital admission following COVID-19 exposure to a group of asymptomatic individuals.

*Added value of this study:* The results indicate that the presence of HLA-DRB1*04:01 might confer protection from the development of respiratory failure following exposure to COVID. Individuals remaining asymptomatic following exposure to COVID are less likely to carry the haplotype DQA1*01:01/DQB1*05:01/DRB1*01:01 compared to the background population. This may indicate a host defence pathway not primarily dependent on an IgG response for clearance of infection. These findings conflict with larger genome wide association studies which compared HLA allelic frequencies of severely unwell patients with the background population.

*Implications of all the available evidence:* The findings could have implications for targeted vaccination regimes as well as helping assess the impact of social restrictions on mortality rates in different populations.

## Background

Since it emerged in Wuhan, China in late 2019, COVID-19 has led to an unprecedented international crisis.(1) At the time of writing, 60 million confirmed cases of infection and over a million deaths have been reported to the World Health Organisation.(2)

The impact of COVID-19 varies markedly, not only between individual patients but also between different populations. At the patient level, it has been shown that males, older patients, those with significant pre-existing medical conditions, and those with an elevated body mass index (BMI) are at increased risk of poor clinical outcomes following exposure. COVID-19 also appears to have disproportionately affected certain ethnic groups and regions of the world.

Analysis of the impact of COVID-19 at a population level is complex, requiring consideration of environmental and socioeconomic factors.(3-5) However, clinical variation in COVID-19 severity and symptomatic presentation may also represent differences in host immunogenetic factors. Through the process of evolution, viruses have exerted selective pressure on humans, meaning that some human populations exhibit marked genetically determined variations in their resistance or susceptibility to different endemic infectious organisms. It is possible that reported regional variations in the impact of COVID may reflect these variations.(6-8)

In this respect, some of the most well described immune features belong to the major histocompatibility complex (MHC). The MHC, located on the short arm of chromosome 6 is one of the most complex genetic systems in the human genome, and comprises the human leukocyte antigen (HLA) genes. The HLA transmembrane proteins encoded by the classical (A, B, C, DR, DQ and DP) HLA genes are principally involved in the antigen presentation, at the cell surface, of small pathogen-derived peptides to T cells, which triggers an immune response.(9) Different HLA alleles present different repertoires of peptide fragments from the invading pathogens, potentially influencing the T cell immune response.(10) Human leukocyte antigen (HLA) gene variants have been implicated in host susceptibility or resistance to diseases such as tuberculosis(11), malaria(8, 12), hepatitis B(13, 14), dengue(15), influenzas(16, 17), SARS(18) and MERS.(19) Bats have evolved highly developed MHCs, and this has been proposed as a reason that they act as a reservoir of coronaviruses, a reservoir from which COVID-19 may have emerged.(20, 21) Furthermore, the expression of HLA genes is known to be influenced by age(22), sex(23) and obesity(24), all identified as key factors in the severity of COVID.

For this study, we investigated whether previously well patients who required admission for treatment of COVID-19 infection have different HLA alleles compared to asymptomatic controls and the background population.

## Methods

### Study Participants and Recruitment

As part of an ethically approved research study (“Do MHC genes play a role in the severity of COVID-19?” IRAS project 283409; REC reference:20/YH/0184) sponsored by North Tees and Hartlepool NHS FT, we compared the classical HLA gene frequencies in two groups: patients admitted with severe COVID infection and hospital staff who remained asymptomatic following exposure to COVID.

### Severe COVID patient group

We recruited 49 patients with severe COVID-19, which was defined as hospitalization with respiratory failure and a confirmed SARS-CoV-2 viral RNA polymerase-chain-reaction (PCR) test from nasopharyngeal swabs, from intensive care units and general wards at two teaching hospitals in the North East of England. Respiratory failure was defined as the requirement for oxygen supplementation and/or mechanical ventilation. Only patients with no significant comorbidities were included. Eligible patients (discharged or inpatient) were identified through consultant review of the medical records. Discharged patients were contacted via phone initially. If the patients expressed a wish to participate, they were sent further correspondence including a patient information sheet providing basic information as to the fundamental reasons for the research study. Subsequently, a pack was sent through the post including a saliva collection kit which was mailed back to the research team.

### Asymptomatic group

We recruited 69 hospital staff members who had tested positive for COVID infection through routine blood antibody testing. Recruitment was carried out through hospital trust email advertising and review of occupational health records to identify staff members who had tested positive for COVID antibodies on blood screening or had tested positive on swab testing. They were contacted by email or telephone and invited to participate in the research. Blood antibody testing was carried out using the Medicines and Healthcare Regulatory Agency (MHRA) approved Abbott (Ilinois, United States) or Roche tests (Basel, Switzerland) which detect immunoglobulin (Ig) G antibodies to Severe Acute Respiratory Syndrome Coronavirus 2 (SARS-CoV-2). For all study participants, patient age, sex, ethnicity and BMI were recorded. Only white British participants were included for this study.

### DNA Sample collection and processing

A combination of ORAcollect OCR-100 buccal swabs and Oragene DNA OG-610 saliva collection kits (both DNA Genotek Inc, Ontario, Canada) were used to collect samples for DNA extraction. DNA was extracted from the swab and saliva samples using the Roche MagnaPure Compact automated platform (Roche Holding AG, Switzerland). DNA was the quantified using the Thermo Fisher Qubit dsDNA BR Assay kit and standardised to 25ng/ul. HLA genotyping was performed using the One Lambda AllType NGS kits (One Lambda, USA), with the Illumina MiSeq platform (Illumina, USA). Full gene sequencing was carried out for HLA-A, -B, -C, -DQA1 and –DPA1 and partial gene sequencing (omission of exon 1) for HLA-DRB1, -DRB345, -DQB1 and –DPB1. HLA genotypes were analysed using One Lambda TypeStream Visual 1.3 software (One Lambda, USA).

### Control groups: Regional and national

Authors of this study are carrying out an ongoing study into the relationship between HLA genotypes and the outcomes of joint replacement surgery. The study involves gene sequencing of patients who have received hip prostheses for end stage degenerative osteoarthritis. The study is being carried out at the same hospital trusts from which the participants of the current study were drawn. 196 patients have undergone full class I and II sequencing and a total of 263 have been sequenced for HLA-DQ typing. These anonymised data, which included patient age and sex, were used as local control data. These data have previously been compared to a larger national data set comprising 8514 National Blood Service and 1958 Birth Cohort controls and were not shown to significantly deviate (supplementary data).

### Statistical analysis

The genotypes for each HLA gene were transformed into dosages of each individual possible allele from within the patient population, where 2 denoted two copies of a given allele, 1 denoted one copy and 0 denoted zero copies. These dosages were then entered as predictor variables in a logistic regression analysis. Three models were tested, the first with the asymptomatic staff versus the severe group, then with asymptomatic staff against background controls, and then with the severe patients as cases against controls. All three models were also tested with sex as an additional covariate and also age plus sex as covariates. Alleles which were found to differ significantly were entered into a binary logistic regression model comparing asymptomatic and severe patients, along with age, sex and BMI.

### Ordinal logistic regression

During the recruitment of asymptomatic staff, we received responses from symptomatic individuals who had tested positive for COVID antibodies who expressed their willingness to participate and provide a sample for DNA analysis. The reported symptoms fit into two broad categories:

“Local symptoms”. These staff members reported symptoms limited to loss of taste and/or smell.

“Systemic symptoms, no hospital treatment”. These staff members reported systemic upset, general malaise, mild fever, arthralgia.

We incorporated the results from these participants into an ordinal logistic regression model using the following ranking scale:

1. Asymptomatic
2. Local symptoms
3. Systemic symptoms, no hospital treatment
4. Systemic symptoms requiring hospital admission

Age, BMI were used as independent variables, along with sex and presence of specific HLA genes identified in the initial comparison between the severe, asymptomatic and control groups.

Global locus-wise association for each HLA gene was performed using UNPHASED v 3.0.13. Haplotypes were estimated for DRB1-DQA1-DQB1 also in UNPHASED.(25)

### Global Distribution of HLA Haplotypes

Given the striking variation in apparent clinical impact of COVID-19 in different geographical locations and in different ethnicities, we accessed the Allele Frequency Net Database(26) to record HLA-DRB1 frequencies in all available gold standard HLA population studies from different nationalities. These were plotted against latitude (supplementary data file 2).

### Role of the funding source

The study was funded by Innovate UK. Innovate UK had no role in the study design, the collection, analysis, interpretation of data, the writing of the report, or in the decision to submit the paper for publication.

## Results

A total of 147 individuals provided samples. Participant and patient details are shown in table 1.

**Table 1:**
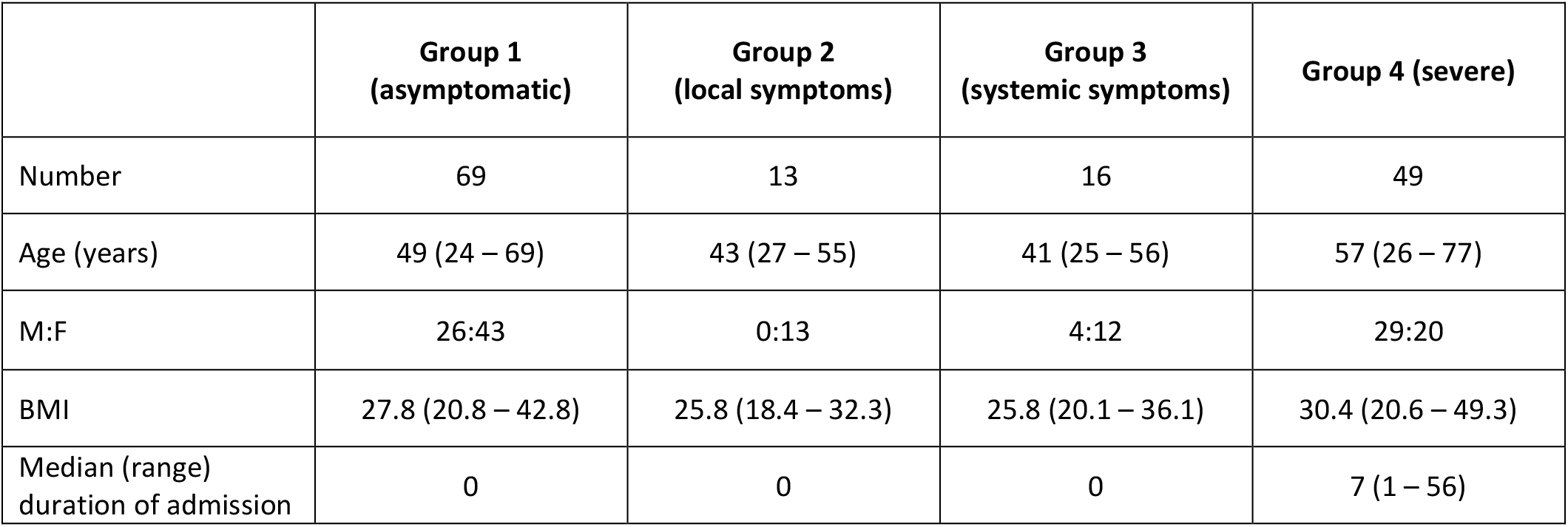
Demographics of the participants in the study groups

### HLA gene frequencies in the asymptomatic staff and severe patient groups

The results, including calculations performed before and after adjustment for age and sex are shown in supplementary data file 1. Locus-wise association between each HLA locus and severe vs asymptomatic COVID-19 showed global association for the DRB1 locus (P<0.008). The alleles which were found to significantly differ between groups are shown in supplementary data file 1. The most robust finding was a significant difference in the allelic frequency of DRB1*04:01 (split equally into DQA1*03:01/DQB1*03:02 and DQA1*03:03/DQB1*03:01) in the severe patient compared to the asymptomatic staff group (5.1% versus 16.6%, p=0.003 after adjustment for age and sex). Carrying this allele appear to protect against severe disease and predispose to an asymptomatic outcome. The frequency of this allele is 11.0% in the study controls and 11.1% in the UK population. DRB1*01:01 (in linkage disequilibrium with DQA1*01:01/DQB1*05:01) was found at a significantly lower frequency in the asymptomatic group (1.4%) compared to the background populations (9.4% in the study controls and 9.7% in the UK population, p=0.007).

### Binary logistic regression

The significant protective effect of HLA-DRB1*04:01 was retained in the binary regression modelling which also incorporated age, sex and BMI. DRB1*04:01 had approximately the same influence on disease severity as patient sex. Age, however, was the dominant variable (table 2).

**Table 2.** Results of the binary logistic regression model.

| Variable | Coefficient | Lower bound<br>(95%) | Upper bound<br>(95%) | Standard<br>error | P value |
| --- | --- | --- | --- | --- | --- |
| Age | 0.575 | 0.280 | 0.871 | 0.151 | <0.001 |
| BMI | 0.465 | 0.187 | 0.742 | 0.142 | 0.001 |
| Sex-M | 0.386 | 0.127 | 0.645 | 0.132 | 0.003 |
| DRB1*04:01<br>carriage | -0.357 | -0.630 | -0.084 | 0.139 | 0.010 |

### Ordinal logistic regression

With all confirmed COVID infected cases (n= 147) included in the ordinal regression model, the same variables were found to be significant. Age (coefficient 0.262, p=0.005), male sex (0.205, p=0.030) and BMI (0.189, p=0.045) were associated with more severe symptoms, and carriage of DRB1*04:01 (−0.326, p=0.001) was associated with less severe symptoms.

### Global distribution of DRB1*04:01

The distribution of DRB1*04:01 showed a positive correlation to latitude (rank correlation = 0.754, p<0.001)(figure 1).

## Discussion

HLA genes are recognised to be important factors in the host response to foreign pathogens. In this regional study, composed of white British participants drawn from a relatively small catchment area, we have identified HLA genes which appear to be associated with the severity of COVID-19 infection.

HLA genes encode for glycoproteins which bind peptides in “peptide binding grooves”, present on the cell membrane. HLA class I molecules, which are present on all nucleated cells, present endogenous peptides to cytotoxic CD8+ T cells. HLA class II molecules are expressed on the surface of antigen-presenting cells (including macrophages, B cells and dendritic cells) and are essential in the presentation of peptides to T-helper CD4+ cells. The variance of individual amino acids within HLA molecules determine the three dimensional structure of the peptide binding groove. The structure of the binding grooves determines which peptides (which can be foreign or auto antigens) are presented at the cell’s surface.(27)

Validated software allows virtual construction of peptide binding grooves encoded by an individual’s HLA genotype.(28) This allows for the calculation of the binding affinity between a particular HLA encoded peptide binding groove and a multitude of naturally occurring peptides. A number of publications have investigated the link between HLA variation and COVID severity using this in silico approach. Some of these studies were combined with epidemiological data(29-31) or results derived from small case control series.(32) There appears to be no clear consensus in the conclusions drawn so far.

The results of the current study indicate that HLA alleles interact with age, sex and BMI to determine clinical outcomes following COVID exposure. One must be cautious in the interpretation of such results, however. HLA genes are by their very nature, highly polymorphic. Due to the number of alleles which are under investigation, the risk of identifying a “significant” allele through chance is high and none of the tests withstood correction for multiple testing. However, the most robust finding appears to be the protection conferred by the presence of DRB1*04:01. Approximately 30% of Caucasians carry DRB1*04, an allele with more than 50 subtypes. Of all the DRB1*04 alleles, *04:01 is the most frequent allele occurring with a frequency of 57% as compared to *0402 which occurs with a frequency of 6.3%.(33)

The various statistical analyses presented in the current paper indicate that while DRB1*04:01 may be protective, other DRB1*04 alleles such as DRB1*04:02 and DRB1*04:05 may be associated with an increase in disease severity. On the face of it, this may seem unusual, yet the profound impact of amino acid substitutions and MHC-restricted T-cell recognition has long been appreciated.(34) For example, DRB1*0402 and *0401 molecules differ in the peptide-binding region by three amino acids yet they have diametrically opposed clinical outcomes: protection from versus susceptibility to rheumatoid disease.(35)

Certain HLA alleles are superior in providing protection by presenting multiple epitopes for the activation of T cells. The evolutionary trade-off is that most autoimmune diseases have been associated with the presence of certain HLA class II molecules. This appears to be the case with HLA-DRB1*04:01, which is associated with rheumatoid disease.(36) Animal studies comparing 04:01 mice with 04:02 show that even though both strains of mice are protected from influenza, only *04:01 mice can generate cross-protective immunity. There is evidence related to influenza to indicate that *04:01 mice show greater proliferation responses and strikingly higher IFN-γ responses in response to vaccination compared to *04:02 mice. Non-antigen specific immune response dictate different signalling pathways and MHC peptide loading occurs in different cell compartments between the closely related HLA genes DRB1*04:01 and DRB1*04:02.(16, 37) This fundamental difference between the two molecules may lead to different peptides being loaded and presented despite the same pathogen exposure. In response to superantigens, *04:01 mice have been shown to produce different and varying amounts of cytokines compared to 04:02 strains.(38)

That the strongest predictor of COVID severity was found in the class II HLA-DR region was not surprising given recently published work.(39, 40) Kachuri et al(3) conducted a comprehensive study including genome-wide and transcriptome-wide association analyses to identify genetic loci associated with IgG antibody response to 16 viruses using serological data from 7924 European ancestry participants in the UK Biobank cohort. Signals in the HLA class II region dominated the antibody response to viruses, with 40 independent loci and 14 independent classical alleles implicated. The strongest associations with seroreactivity were identified within the DRβ1 locus.

These results were substantiated by Hammer et al(41) who carried out a cross-pathogen, genome-wide investigation of the role of host genetics in modulating the individual IgG response to common viral antigens. Hammer et al found that individuals carrying DRB1*15:01 were more likely to have detectable levels of anti-influenza A IgG, whereas the presence of DRB1*01:01 was associated with seronegativity.

Consistent with data from the biobank studies, we found an increased frequency of DRB1*15:01 in both the asymptomatic and severe groups when compared to the background controls. This finding did not achieve significance however in our study. We believe this may have been due to our study protocol, which we designed primarily to identify differences between severe and asymptomatic groups. However, in a study of HLA 99 Italian patients admitted for COVID-19 infection, (42) Novelli et al did observe significantly increased frequencies of DRB1*15:01 and DQB1*06:02 (alleles which are recognised to be in strong linkage disequilibrium). The authors noted that their findings conflicted with a larger study by Ellinghaus et al(32), who conducted a genome wide association study involving 1980 patients with COVID-19 and severe disease (defined as respiratory failure) at seven hospitals in Italy and Spain. Ellinghaus et al found no SNP association signals at the HLA complex that met even the significance threshold of suggestive association with either COVID-19 or disease severity.

In the Ellinghaus study, the median ages of the patients recruited from seven centres varied from 64 to 69 years with interquartile values ranging from 54 to 79 years. These patients were significantly older and frequently exhibited other medical comorbidities and the hospitals were separated by over 1500 kilometres. In the current study, the median age of the admitted patients was 59 years and all hospitals and the catchment for the control group, were separated by a maximum distance of 80 kilometres.

Other issues to consider may include patient exposure to different COVID strains(30, 43) and environmental factors. The effects of climate and seasonal changes on virus transmission have been well documented. Multiple studies emanating from several countries indicate that humidity and temperature changes are critical factors in viral outbreaks.(3, 44, 45) The evidence for sunlight exposure and vitamin D is less convincing.(46) However, vitamin D has long been known to exhibit immunomodulatory effects and there is evidence for vitamin D action on MHC class II gene expression.(47) Ultraviolet radiation dependent metabolism provides the major source of vitamin D in humans. At higher latitudes, solar radiation is generally insufficient to act as the primary source during the winter however.(47, 48) As a result, vitamin D deficiency is associated with numerous autoimmune and infectious diseases the distribution of which, in some cases, mirrors that of reduced UVB exposure.(48) A number of the genes linked to COVID severity in the current study have been shown to exhibit vitamin D receptors.(49, 50) There is a large amount of data linking vitamin D, multiple sclerosis and DRB1*15, but for the other HLA genes it is less clear(51, 52). Interestingly, the frequency of genes modulated by vitamin D increases with latitude (supplementary data). We speculate that the role of vitamin D may be as a suppressive agent, regulating the immune system on a seasonal basis to strike a balance between clearance of infection versus the susceptibility to autoimmune disease.

The haplotype DQA1*01:01/DQB1*05:01/DRB1*01:01 was found at a significantly lower frequency in the asymptomatic group when compared to the background population. The lower frequency of this haplotype in the severe patients suggest that response to COVID in these individuals may not be primarily dependent upon IgG antibody production. However, there appeared to be significant sex interaction, with a haplotype frequency in the severe female group of 12.5% compared to 3.4% in the males. Interestingly, in our ongoing investigation into responses to orthopaedic implants, DRB1*01:01 was significantly associated with a lower probability of T and B cell infiltration into periprosthetic tissues in female patients and this was one of the factors which prompted the current investigation. Reported male to female COVID mortality ratios vary widely across the globe. Published data indicate that this variation may be associated with variation in DRB1*01:01 frequency in different populations (supplementary data 2).

## Summary

We have provided evidence to show that HLA alleles interact with patient factors to influence susceptibility to developing severe complications from COVID-19. HLA alleles associated with asymptomatic carriage of the disease are relatively common in Caucasian populations living at higher latitudes.

## Data Availability

Authors agree to make data and materials supporting the results or analyses presented in their paper available upon reasonable request. It is up to the author to determine whether a request is reasonable.

## Author contributions

All authors had full access to all the data in the study and have accepted responsibility to submit for consideration of publication. All authors took part in the design of the study, data collection and the drafting and final approval of the manuscript.

## Declaration of interests

Based on these results DJL has filed a patent “Biomarkers for Covid-19 Symptom Severity”.

### Supplementary data 2: Relationship of HLA-DR alleleic frequencies and latitude

Common DRB1 alleles and their relationship to latitude. Significant positive correlations to latitude shown in green, negative in pink.

| DRB1 allele | Rank correlation to latitude | P value |
| --- | --- | --- |
| 01:01 | 0.818 | <0.001 |
| 03:01 | 0.740 | <0.001 |
| 04:01 | 0.754 | <0.001 |
| 04:02 | 0.187 | 0.390 |
| 04:03 | -0.563 | 0.006 |
| 04:04 | 0.628 | 0.002 |
| 04:05 | -0.115 | 0.592 |
| 04:06 | -0.124 | 0.564 |
| 04:07 | 0.173 | 0.426 |
| 04:08 | 0.485 | 0.023 |
| 07:01 | 0.520 | 0.010 |
| 08:01 | -0.115 | 0.601 |
| 09:01 | -0.473 | 0.021 |
| 10:01 | -0.353 | 0.092 |
| 11:01 | -0.151 | 0.479 |
| 11:04 | 0.181 | 0.407 |
| 12:01 | -0.380 | 0.068 |
| 13:01 | 0.726 | <0.001 |
| 13:02 | 0.349 | 0.094 |
| 14:01 | -0.611 | 0.002 |
| 15:01 | 0.778 | <0.001 |
| 15:02 | -0.720 | <0.001 |
| 16:01 | 0.168 | 0.441 |
| 16:02 | -0.667 | 0.001 |

### HLA-DRB1*01:01, male to female mortality and latitude

As reported in the main text, DRB1*01:01 (in linkage disequilibrium with DQA1*01:01/DQB1*05:01) was found at a significantly lower frequency in the asymptomatic group (1.4%) compared to the background populations (9.4% in the study controls and 9.7% in the UK population, p=0.007). This could indicate that patients with this allele are less likely to be asymptomatic, or that patients with this allele do not depend on an IgG mediated response to COVID-19 infection. However, there appeared to be significant sex interaction, with a haplotype frequency in the severe female group of 12.5% compared to 3.4% in the males. To investigate this further, we examined published data to see if we could identify global trends which could be consistent with our results.

We accessed the Allele Frequency Net Database to record HLA-DRB1*01:01 frequencies in all available gold standard HLA population studies from different nationalities and compared them to national male to female COVID mortality rates, as reported by Global Health 5050 (https://globalhealth5050.org/the-sex-gender-and-covid-19-project/dataset) (supplementary data 2 figure 1).

In order to identify any significant interaction with other variables we included the following data in a subsequent regression model: latitude of population, mean BMI (from “Global status report on noncommunicable diseases 2014”, World Health Organization report published 2018), mean life expectancy (from the UN Human Development Report 2019) and GDP per capita (from the “World Economic Outlook - GDP per capita”, International Monetary Fund, October 2020).

**Supplementary data 2 figure 1.**
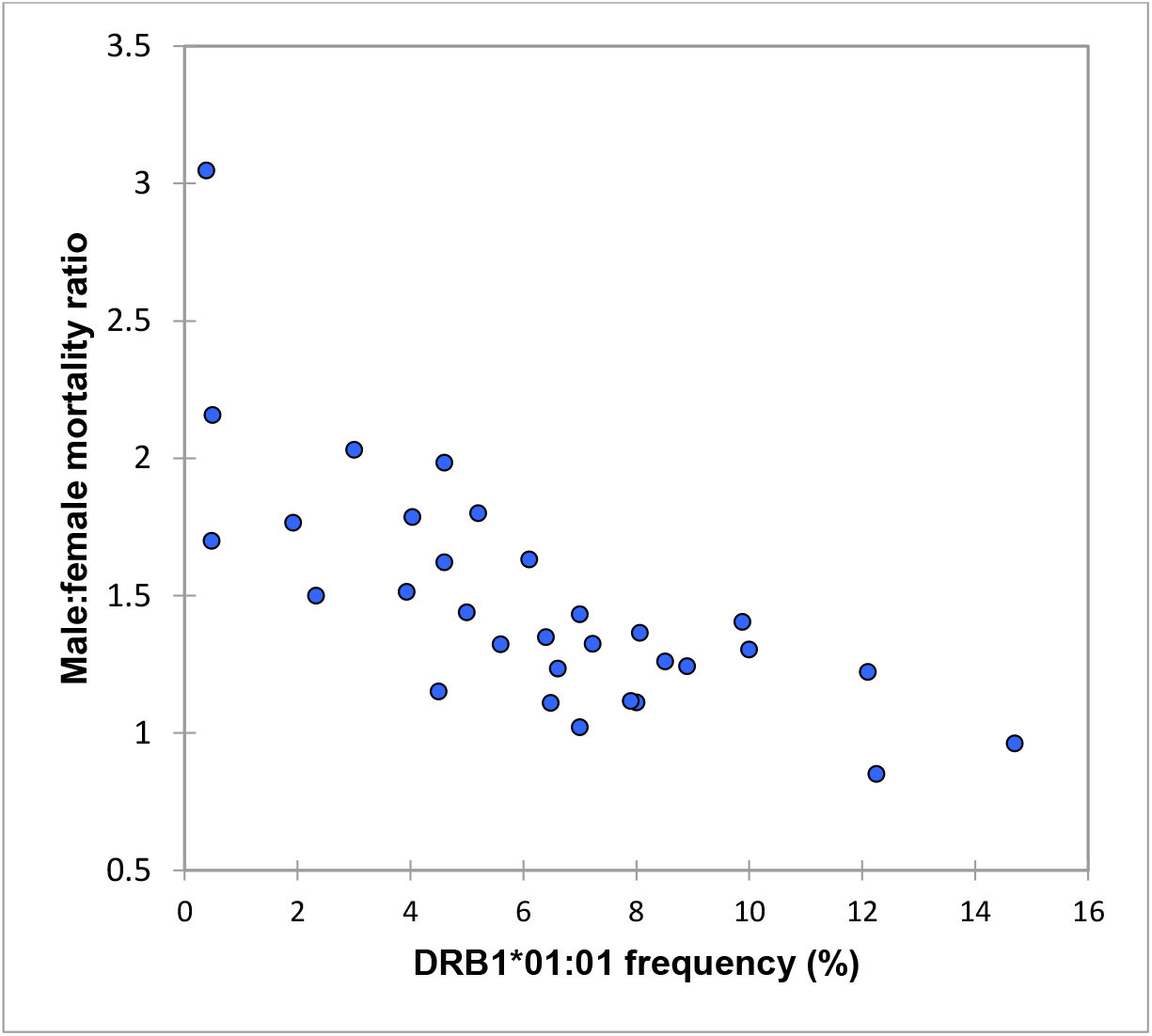
Relationship between male to female mortality ratio and HLA-DRB1*01:01 frequencies in 31 countries across the world (rank correlation = -0.760, p<0.001).

The only significant variables found to influence male to female mortality ratios were latitude and DRB1*01:01 frequency. These two variables returned an R2 value of 64% (p <0.001).

**Supplementary data 2 table 1.** Results of regression modelling to explain variation in male to female mortality ratios.

| Source | Coefficient | Standard error | P value | Lower bound (95%) | Upper bound (95%) |
| --- | --- | --- | --- | --- | --- |
| Latitude (degrees) | -0.469 | 0.162 | **0.007** | -0.802 | -0.137 |
| DRB1*01:01 frequency (%) | -0.395 | 0.162 | **0.021** | -0.727 | -0.063 |

**Supplementary data 2 figure 2.**
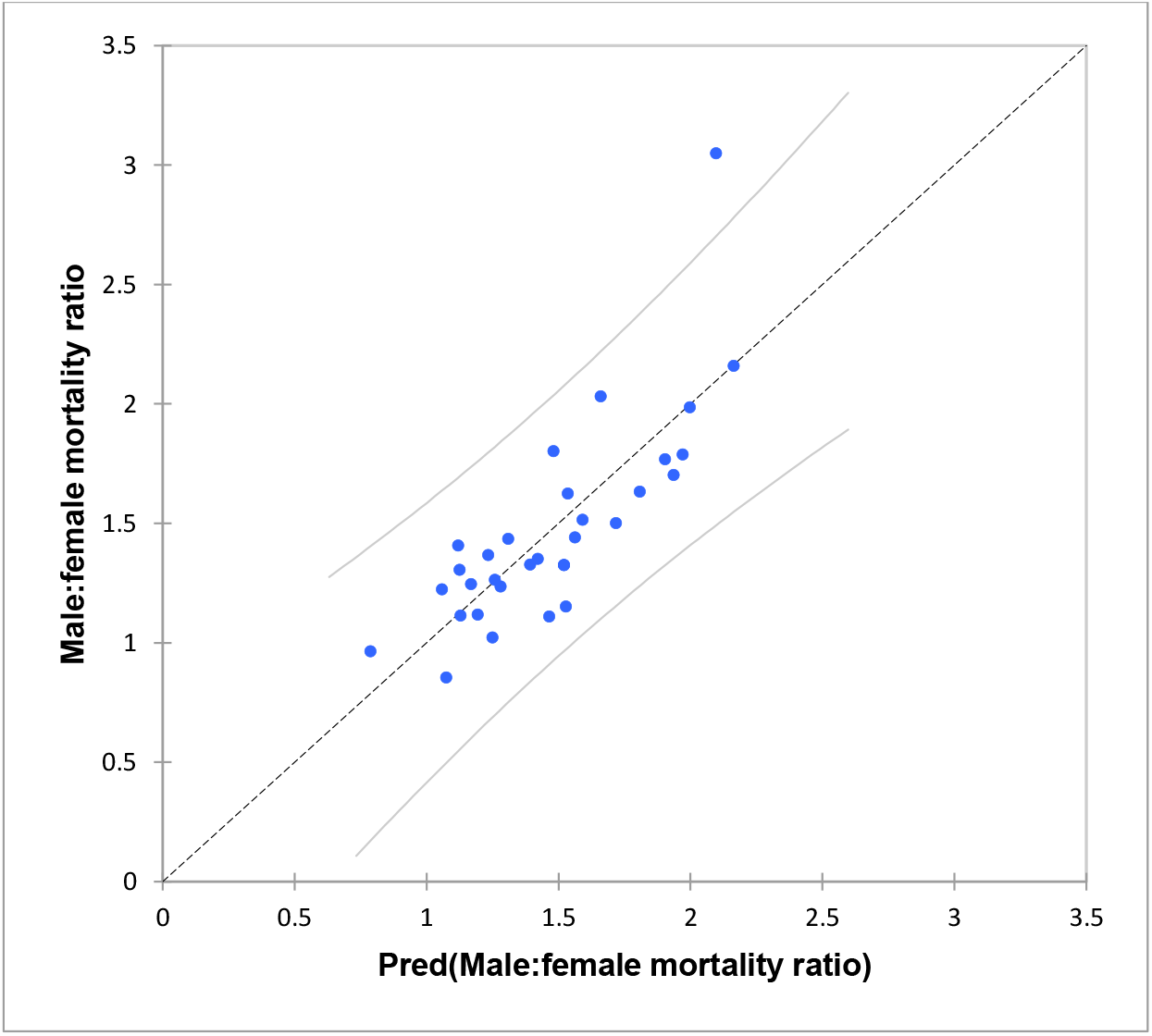
Results of regression modelling incorporating latitude and DRB1*01:01 frequency.

## Notes

### Funding Statement

Funding was received from Innovate UK and ExplantLab.

### Author Declarations

IRAS project 283409: "Do MHC genes play a role in the severity of COVID-19?". REC reference:20/YH/0184) Sponsored by North Tees and Hartlepool NHS FT.

